# Indirect Benefits of Seasonal Malaria Chemoprevention for Non-Malarial Pediatric Infections and Routine Antibiotic Use in Real-World Programmatic Settings: A Pre-Post Study Using Positive and Negative Controls

**DOI:** 10.1101/2025.05.08.25327228

**Authors:** Elisabeth A. Gebreegziabher, Mamadou Ouattara, Mamadou Bountogo, Boubacar Coulibaly, Valentin Boudo, Thierry Ouedraogo, Elodie Lebas, Huiyu Hu, Kieran S. O’Brien, Michelle S. Hsiang, David V. Glidden, Benjamin F. Arnold, Thomas M. Lietman, Ali Sié, Catherine E. Oldenburg

## Abstract

**Objective:** To assess the benefits of Seasonal Malaria Chemoprevention (SMC), the monthly administration of sulfadoxine-pyrimethamine and amodiaquine during the high malaria season, beyond malaria prevention in real-world program settings.

**Methods:** We conducted a pre-post comparison of non-malarial diagnoses (pneumonia, diarrhea, acute malnutrition) and antibiotic prescription rates during SMC administration weeks versus a three-week post-intervention period in rural Burkina Faso. Data came from clinic surveillance at 51 health facilities, a population-based census, and National Malaria Control Program data on SMC timing. Poisson regression models with person-weeks as an offset and standard errors clustered by health post estimated changes in rates. Interaction terms assessed variation across SMC cycles. Positive (malaria diagnoses, antimalarial prescriptions) and negative (injury) control outcomes were used to evaluate potential unmeasured confounding.

**Results:** Compared to administration weeks, modest declines were observed in pneumonia, diarrhea, and acute malnutrition diagnoses, as well as in antibiotic prescription rates during the post-SMC period. Absolute reductions were 0.7 (95% CI: 0.3–1.0), 0.2 (95% CI: 0.1–0.4), 0.05 (95% CI: 0.001–0.09), and 0.90 (95% CI: 0.4–1.4) per 1,000 person-weeks, respectively. Positive control outcomes also declined, with malaria diagnoses and antimalarial prescriptions decreasing by 3.7 (95% CI: 2.6–4.8) and 3.6 (95% CI: 2.5–4.7) per 1,000 person-weeks. Injury rates (negative control) remained stable (0.02; 95% CI: −0.03 to 0.07). Reductions varied across SMC cycles and were most pronounced following the final round.

**Conclusion:** SMC may have additional benefits beyond malaria prevention, including reductions in common pediatric infections and subsequent routine antibiotic use.

## Introduction

Infectious diseases, particularly acute diarrheal and respiratory infections, and malaria, are the leading causes of illness and death in children in low– and middle-income countries.^1^ One strategy to prevent malaria infections and subsequent child mortality is the administration of seasonal malaria chemoprevention (SMC) for children under five. SMC involves administration of sulfadoxine-pyrimethamine and amodiaquine (SP-AQ) during the rainy season in areas with highly seasonal malaria transmission and regions where *Plasmodium falciparum* strains are sensitive to this antimalarial.^2^ In most regions, SMC is administered in four or five monthly cycles, beginning July of each year.

In 2012, the World Health Organization recommended the adoption of SMC, and since then, several African countries have implemented, with over 50 million children receiving it in 2023.^3–5^ Recent guidelines have introduced more flexibility, allowing for adjustments in age groups, dosages, and treatment frequency based on local conditions.^6^ Evidence from randomized controlled trials (RCTs) and observational studies show that SMC is an effective malaria control strategy.^7–9^

Although SMC has been shown to prevent clinical malaria and reduce all-cause mortality,^10,11^ its effect on non-malarial infections is not clear. While SMC primarily targets malaria, the SP component has weak antimicrobial/antibiotic properties that may help prevent or treat other infections.^12,13^ It remains unclear whether there are additional mechanisms through which SMC might lower all-cause mortality beyond its effects on malaria. Additionally, malaria may increase susceptibility to other infections through indirect effects on the immune system,^14^ anemia,^15^ and nutrition/growth^16^. Therefore, by reducing malaria morbidity and susceptibility to other infections, malaria control efforts such as SMC may have potential to offer broader health benefits for children beyond those directly linked to malaria reduction.

Previous research suggests that clearance of chronic malaria was associated with lower overall illness, decreased antibiotic use and improved immunocompetence against non-malaria infections among children.^17^ Effective malaria control interventions may reduce other infections and routine antibiotic use,^17^ potentially contributing to improvements in overall health and a reduction in selection for antibiotic resistance from routine use. Therefore, examining the impact of SMC on overall infection burden, common infections such as pneumonia and diarrhea, and subsequent antibiotic use, can help determine whether it reduces non-malarial illnesses, offers broader health benefits beyond malaria, and potentially help in designing integrated health interventions.

Furthermore, previous research shows variability in the effectiveness of SMC across cycles, with the greatest benefit against malaria observed in the fourth and final cycle.^18^ Understanding whether any indirect benefits SMC may have vary over the course of administration may help health programs optimize and integrate malaria interventions with broader infectious disease control efforts.

To explore this, we assessed the short-term effects of SMC on non-malarial outcomes in the weeks following treatment by analyzing routine surveillance data on diagnoses for children under five from primary healthcare facilities in Nouna, Burkina Faso. The objectives of this study were to 1) to determine whether and how non-malarial outcomes such as pneumonia, diarrhea, and acute malnutrition among children under five change following SMC administration; 2) compare overall antibiotic use at the population level in the weeks before and after SMC; and 3) assess whether changes, if any, in non-malarial outcomes following SMC, or indirect health benefits, vary by month of administration. We hypothesized that SMC could have indirect benefits and help lower non-malarial diagnoses and antibiotic prescription rates.

## Methods

### Study Setting and Population

Health posts (n=51) in the rural northwestern district of Nouna in Burkina Faso, including both the Nouna Health and Demographic Surveillance Site (HDSS),^19^ and the surrounding non-HDSS area^20^ were included in the analysis. In Burkina Faso, children under five years of age receive free care at Centres de Santé et de Promotion Sociale (CSPS), which are government-run, nurse-led primary healthcare facilities offering basic preventive and treatment services, including antenatal care, postnatal care, and vaccinations for children. These centers are often the first line of care and provide referrals for more advanced cases. The median rate of healthcare visits in the villages served by these health posts in 2020 was 6.7 per 100 child-months with the most common reasons for visits being pneumonia (37.5%), malaria (25.1%) and diarrhea (9.1%) infections.^21^ In this analysis, we included data on diagnoses from 2020 and 2021 for children aged 1-59 months, as reported by the health posts in the study area.

The study was reviewed and approved by the Institutional Review Boards at the University of California, San Francisco and the Comité National d’Ethique pour la Recherche (National Ethics Committee of Burkina Faso) in Ouagadougou, Burkina Faso.

### Data Sources and Measures

We used three data sources for this analysis: 1) primary healthcare surveillance, 2) census data from the Community Health with Azithromycin Trial (CHAT) (ClinicalTrials.gov Identifier: NCT03676764),^22^ and 3) Burkina Faso’s National Malaria Control Program (NMCP) data.

#### Primary Healthcare Surveillance

Passive surveillance was conducted at each CSPS (health post) in the study area. Each visit by a child under five was recorded, including the reason for the visit (e.g., fever, diarrhea, malnutrition, etc.), the village of residence, the child’s age and sex, the diagnosis (e.g., malaria, pneumonia, etc.), the treatment provided (e.g., antibiotics, antimalarial agents, etc.), and the timing of the visit (e.g., first versus follow-up visit). Data were routinely collected by facilities on paper forms and were entered into an electronic database. Clinic visit counts for malaria, pneumonia, diarrhea, acute malnutrition and injury cases were used to determine diagnosis rates for their respective outcomes. Malaria was typically diagnosed using a rapid diagnostic test (RDT) and based on symptoms during stockouts.^23^ Other diagnoses were determined based on the WHO community management of childhood illness algorithms, but the accuracy of these diagnoses was not captured.^23^ Acute malnutrition included both moderate and severe forms of acute malnutrition, which were combined due to the low number of cases. Injuries included various types, such as bites, burns, and wounds. Treatments prescribed by healthcare providers were grouped by type at entry, and the counts of antibiotics and antimalarials were analyzed. We restricted the surveillance data to villages in Nouna where the census was conducted and to the year 2020 and 2021, for which SMC administration data were obtained.

#### CHAT Trial Data

CHAT was a cluster RCT of 285 villages, designed to evaluate the efficacy of mass and targeted azithromycin strategies for reducing child mortality.^22^ It was conducted for 3 years (2019-2023). The census performed for the CHAT trial was used to generate the population denominator for this study. CHAT’s methods, including the setting and size of the study, have been described in detail previously.^20,22^ In this trial, a census was conducted every 6 months documenting births, deaths, pregnancies, and migration. The 2020 and 2021 census data were used as denominator to derive the number of children in each health post, which was then used to calculate diagnoses rates.

<u>NMCP Data:</u> these data were used to obtain SMC administration dates and the corresponding epidemiological (epi) weeks for each round of treatment in 2020 and 2021. Epi week or a CDC week begins on a Sunday and ends on Saturday and is a standardized method of counting weeks to allow comparison of data year after year.^24^ In 2020, SMC was administered on July 13-16 (epi-week 29), August 12 – 15 (epi-week 33), September 11-14 (epi-week 37) and October 10-13 (epi-week 41). In 2021, it was administered on July 6-9 (epi-week 27), August 3 – 6 (epi-week 31), August 31-September 2 (epi-week 35) and September 28-October 1 (epi-week 39). During each round, SMC was administered to the entire study area during the same epi week over a 3-4 day period.

### Study Design and Statistical Analysis Methods

We used a one-group pretest-posttest design with multiple follow-up measurements,^25^ that were then combined into a post-treatment period. We compared diagnoses and treatment prescription rates during the administration weeks with rates in the three weeks following administration. The three weeks were combined as post-intervention weeks since SMC is conducted every 4 weeks, and the effect has also been shown to last for only the few weeks after administration.^26,27^ We used a conceptual framework (Figure 1) to show the relationships between variables and how SMC could be related to changes in other infections and antibiotic prescription rates. However, in the analyses, we only adjusted for the cycle (month of administration), a time-related factor, as some of these diagnoses are seasonal, with the month potentially influencing these rates. Since our assessment examines rates in the same population over time, it minimizes bias from time-invariant factors at the population level, which is one of the benefits of pre-post comparisons.^28^

**Figure 1.**
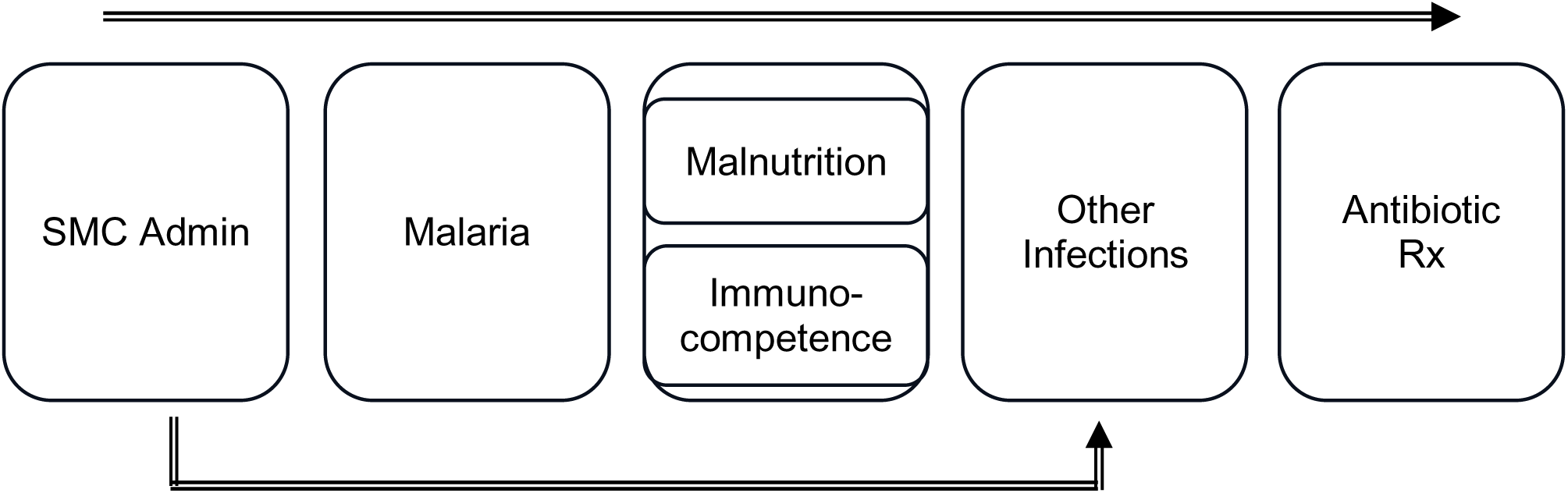
– Conceptual Framework for SMC, Non-Malarial Outcomes, and Antibiotic Prescription Rates. **Note**: This conceptual framework illustrates how SMC administration is hypothesized to affect non-malarial diagnoses and antibiotic prescriptions, though it does not depict every relationship between these factors/variables.

To evaluate potential influence of unmeasured confounding from other factors that may change over time, we used both negative and positive control outcomes.^29,30^ Specifically, since we did not have a control group to account for changes in the absence of intervention, we used these control outcomes to help identify the presence of unmeasured confounding or other factors influencing outcomes. We used malaria as the positive control outcome, as it is the outcome for which SMC is intended and has been proven effective.^7,8^ Injury rate was used as a negative control outcome as it would share the same potential sources of bias (given that these were reported by the same facilities using the same process) and is an outcome that cannot be related to treatment (SMC).^29,30^ Additionally, we examined how antimalarial prescription rates in 2020 and 2021 changed following SMC administration (which were expected to change with malaria), to serve as a positive control outcome for antibiotic prescription rates. We used methods similar to those used in the main analyses described below to examine changes in control outcomes following SMC administration, compared to administration weeks.

### Statistical Analysis

Diagnoses for each year (2020 and 2021) were aggregated to counts by epi week for each of the 51 health posts (CSPS) in the study area. Person-weeks were calculated using the total number of children in each CSPS for each year based on the census data. The surveillance and census data at the health post level were joined by CSPS. For each CSPS, weekly diagnoses counts were included in a longitudinal format. We estimated incidence rates and 95% confidence intervals for several epi weeks before and after administration to describe diagnoses and treatment prescription trends during and after SMC distribution in 2020 and 2021.

For the pre-post comparison, the administration weeks (8 epi-weeks across the 51 health posts) served as reference against which the rates for the post administration period (the three weeks after (24 epi-weeks)) were compared. Approximately 10% of the epi weeks across all health posts did not have recorded counts of diagnoses and treatments. Given the limited amount of missing data, we conducted a complete-case analysis.

We used a Poisson regression model with person-weeks used as an offset and standard errors clustered by health post. We calculated marginal effects, expressed as incidence rate ratios (IRR) and incidence rate differences (IRD) across all health posts. To examine whether the effect of SMC varied across cycles, we included an interaction term between post-SMC period and SMC-cycle in the models. Results were considered statistically significant at an alpha level of p < 0.05. To adjust for multiple comparisons, we used the Romano-Wolf method with 1000 repetitions, which uses bootstrap resampling to control for the familywise error rate.^31^

Since the benefit of SMC for malaria has been shown to decline in the weeks following administration,^27,32^ as a sensitivity analysis, we examined how rates of diagnoses and treatment prescriptions following SMC changed in the first, second and third week post administration.

SAS 9.4 (SAS Institute, Cary, NC) was used for data cleaning and for generating the dataset for analysis. Stata version 14.2 (StataCorp, College Station, TX) was used for all other analyses.

## Results

The characteristics of the health posts included in the analysis are shown in Table 1. The average number of children 1 to 59 months of age across the 51 health post catchment areas was 2,246. The average age was 38.3 months (3.2 years), and approximately 50% of the children were male. Among diagnoses, malaria, pneumonia and diarrhea were most frequent with average rates of 6.0 (95% CI: 5.2–6.9), 4.3 (95% CI: 3.5–5.2), and 1.1 (95% CI: 0.8–1.4) cases per 1,000 person-weeks, respectively. Acute malnutrition and injury were rare with average rates of 0.12 (95% CI: 0.07 –0.18) and 0.16 (95% CI: 0.13–0.19) cases per 1,000 person-weeks. Anti-malarial prescription rates closely mirrored that of malaria with an average rate of 6.0 (95% CI: 5.1 – 6.9) prescriptions per 1,000 person-weeks, while antibiotic prescription rates were slightly higher, at 6.9 (95% CI: 5.7 – 8.0) prescriptions per 1,000 person-weeks. By quarter of year, malaria and pneumonia diagnoses as well as treatment prescription rates were highest in the fourth quarter (Oct-Dec) while diarrhea rates were relatively higher in the first (Jan-Mar) and third (July-Sep) quarter of the year.

**Table 1.**
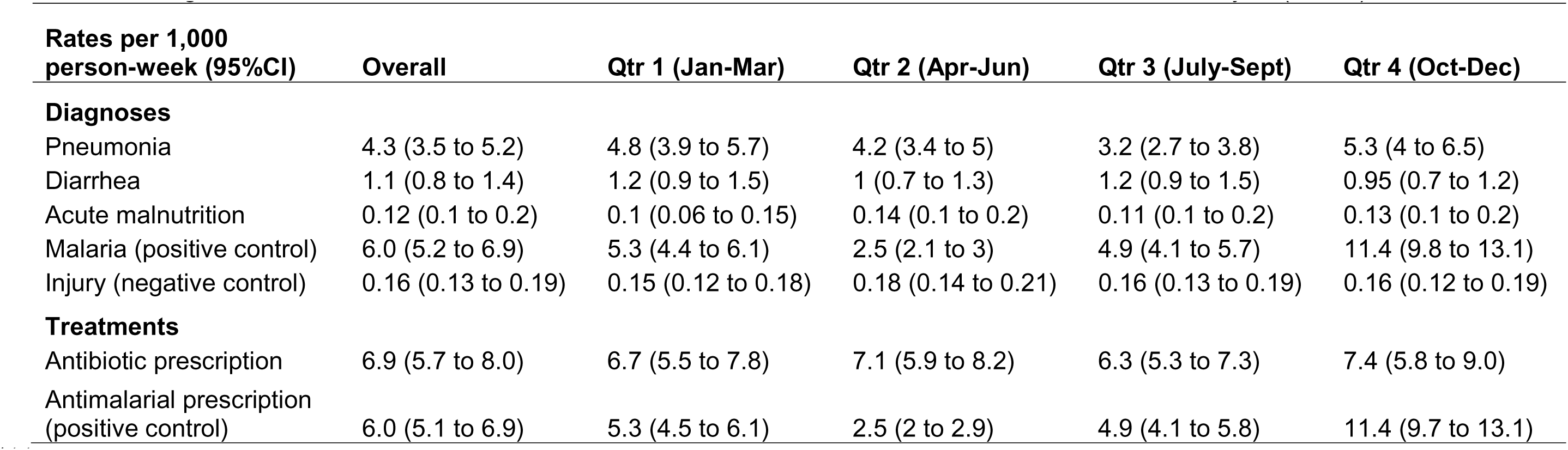
– Diagnoses and treatment rates in CSPS/Health Posts in Nouna, Burkina Faso included in the Analysis (n = 51), 2020 and 2021.

### Changes in Diagnosis and Treatment following SMC

Figure 2 shows the rates of diagnoses and treatment prescriptions in 2020 and 2021 during the multiple weeks before and after SMC administration. Pneumonia diagnosis and antibiotic prescription rates showed some seasonality, with relatively higher rates toward the end of the year (after the administration of the fourth round of SMC). In contrast, acute malnutrition rates remained relatively constant in the weeks before, during, and after the SMC, while diarrhea rates were slightly higher around the weeks of SMC administration, with small fluctuations in trends (Figure 2).

**Figure 2.**
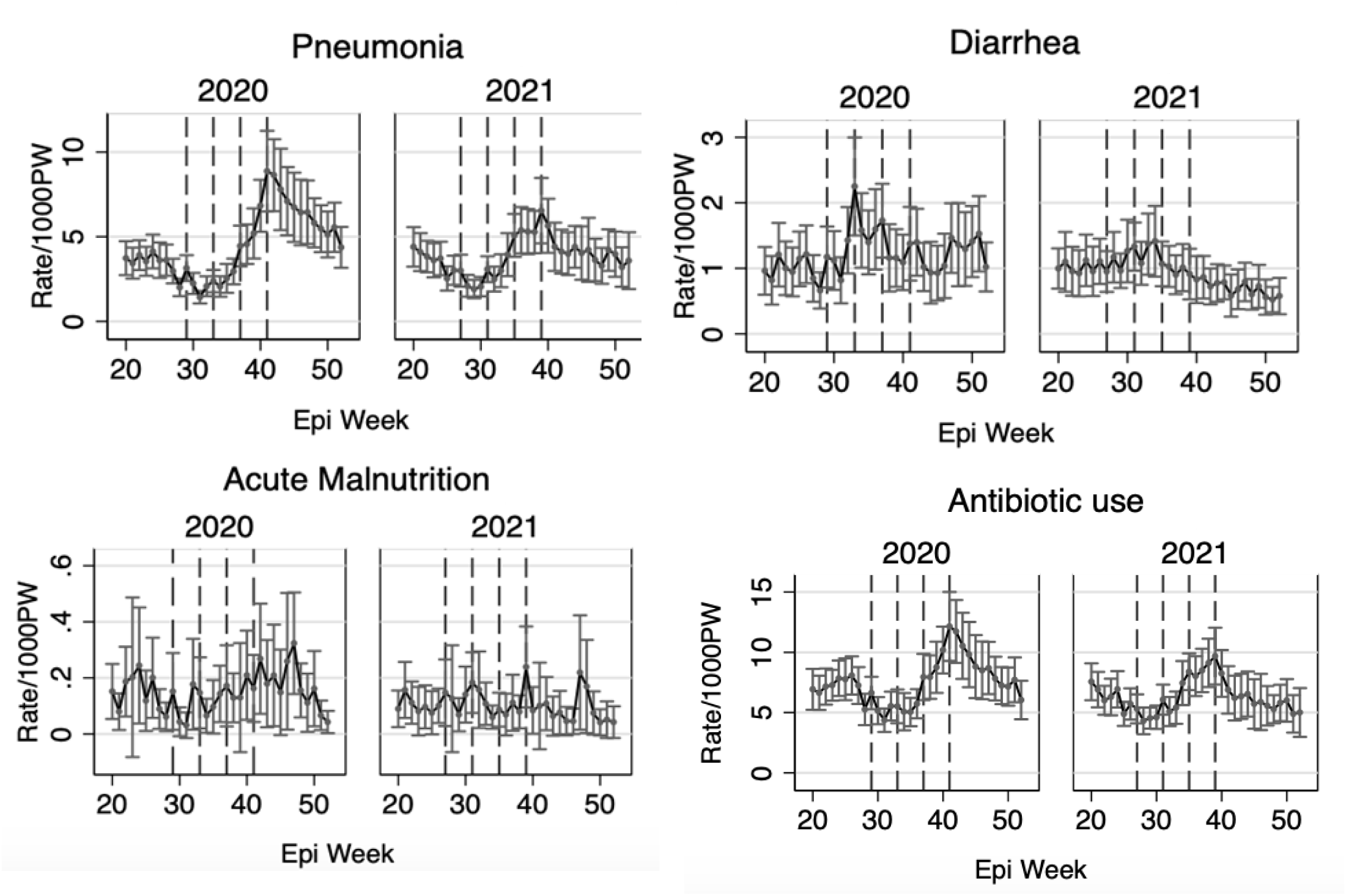
– Diagnosis Rates and Treatment Prescription Rates in 2020 and 2021 in the Context of SMC Administration. Dashed vertical lines mark the four SMC administrations in each year of surveillance

For the control outcomes, malaria and antimalarial prescription rates showed high seasonality, with higher rates toward the end of the year, while injury rates remained relatively constant throughout the weeks (Supplementary Figure 1).

In the pre-post comparison, compared to the administration weeks, the rates of pneumonia, diarrhea, and acute malnutrition declined by 14% (95% CI: 7%–21%), 17% (95% CI: 7%–26%), and 29% (95% CI: 3%–49%), respectively (Table 2). Following SMC administration, antibiotic prescription rates declined by 12% (95% CI: 6%–17%).

**Table 2.** – Incidence rate, rate ratio and rate difference of outcomes on SMC administration weeks and the three weeks post administration.

| Pre and Post SMC administration | Incidence rate per 1,000 person-week (95%CI) | Incidence rate ratio (95%CI) | Incidence Rate Difference per 1,000 person-week (95%CI) | P value | Romano-wolf adjusted p value |
| --- | --- | --- | --- | --- | --- |
| **Pneumonia** |  |  |  |  |  |
| Admin weeks | 4.7 (3.8 to 5.6) | Ref | Ref |  |  |
| Post admin weeks | 4.1 (3.3 to 4.8) | 0.86 (0.79 to 0.93) | -0.7 (-1.0 to -0.3) | 0.000 | 0.003 |
| **Diarrhea** |  |  |  |  |  |
| Admin weeks | 1.4 (1 to 1.7) | Ref | Ref |  |  |
| Post admin weeks | 1.1 (0.8 to 1.4) | 0.83 (0.74 to 0.93) | -0.2 (-0.4 to -0.1) | 0.002 | 0.003 |
| **Acute Malnutrition** |  |  |  |  |  |
| Admin weeks | 0.17 (0.09 to 0.24) | Ref | Ref |  |  |
| Post admin weeks | 0.12 (0.05 to 0.18) | 0.71 (0.51 to 0.97) | -0.05 (-0.09 to 0.0) | 0.034 | 0.040 |
| **Control diagnoses outcomes** |  |  |  |  |  |
| **Malaria (positive control)** |  |  |  |  |  |
| Admin weeks | 9.8 (8.1 to 11.5) | Ref | Ref |  |  |
| Post admin weeks | 6.1 (5.1 to 7) | 0.62 (0.56 to 0.69) | -3.7 (-4.8 to -2.6) | 0.000 | 0.003 |
| **Injury (negative control)** |  |  |  |  |  |
| Admin weeks | 0.17 (0.12 to 0.23) | Ref | Ref |  |  |
| Post admin weeks | 0.15 (0.12 to 0.19) | 0.89 (0.67 to 1.19) | -0.02 (-0.07 to 0.03) | 0.422 | 0.232 |
| **Treatments** |  |  |  |  |  |
| **Antibiotic use** |  |  |  |  |  |
| Admin weeks | 7.9 (6.6 to 9.2) | Ref | Ref |  |  |
| Post admin weeks | 6.9 (5.8 to 8.1) | 0.88 (0.83 to 0.94) | -0.9 (-1.4 to -0.40) | 0.000 | 0.003 |
| **Antimalarial use (positive control)** |  |  |  |  |  |
| Admin weeks | 9.7 (8 to 11.4) | Ref | Ref |  |  |
| Post admin weeks | 6.1 (5.1 to 7.1) | 0.63 (0.56 to 0.69) | -3.6 (-4.7 to -2.5) | 0.000 | 0.003 |

For control outcomes, malaria rates (the positive control outcome) declined by 38% (95% CI: 31% to 44%), while injury rates (the negative control outcome) remained unaffected during the post-SMC administration period, as expected (Table 2). Antimalarial prescription rates (the positive control outcome) decreased by 37% (95%CI: 31% to 44%), mirroring changes in malaria rates.

The statistical significance of the findings remained unchanged after adjusting for multiple comparisons using Romano-Wolf P-values, with all outcomes except injury rates remaining statistically significant (Table 2).

### Variation in Changes in Outcomes by Month of Administration

For the comparison of changes across SMC cycles, the change in rates following SMC administration varied by month (cycle) for pneumonia, diarrhea, and antibiotic prescriptions (P values for the global test of interaction= 0.000, 0.047, and 0.002, respectively; Figure 3, Supplementary Table 1). Among the four cycles, the change in rates following SMC administration was most pronounced in October for pneumonia and antibiotic prescriptions, and in August for diarrhea. No significant variation was observed for acute malnutrition.

**Figure 3.**
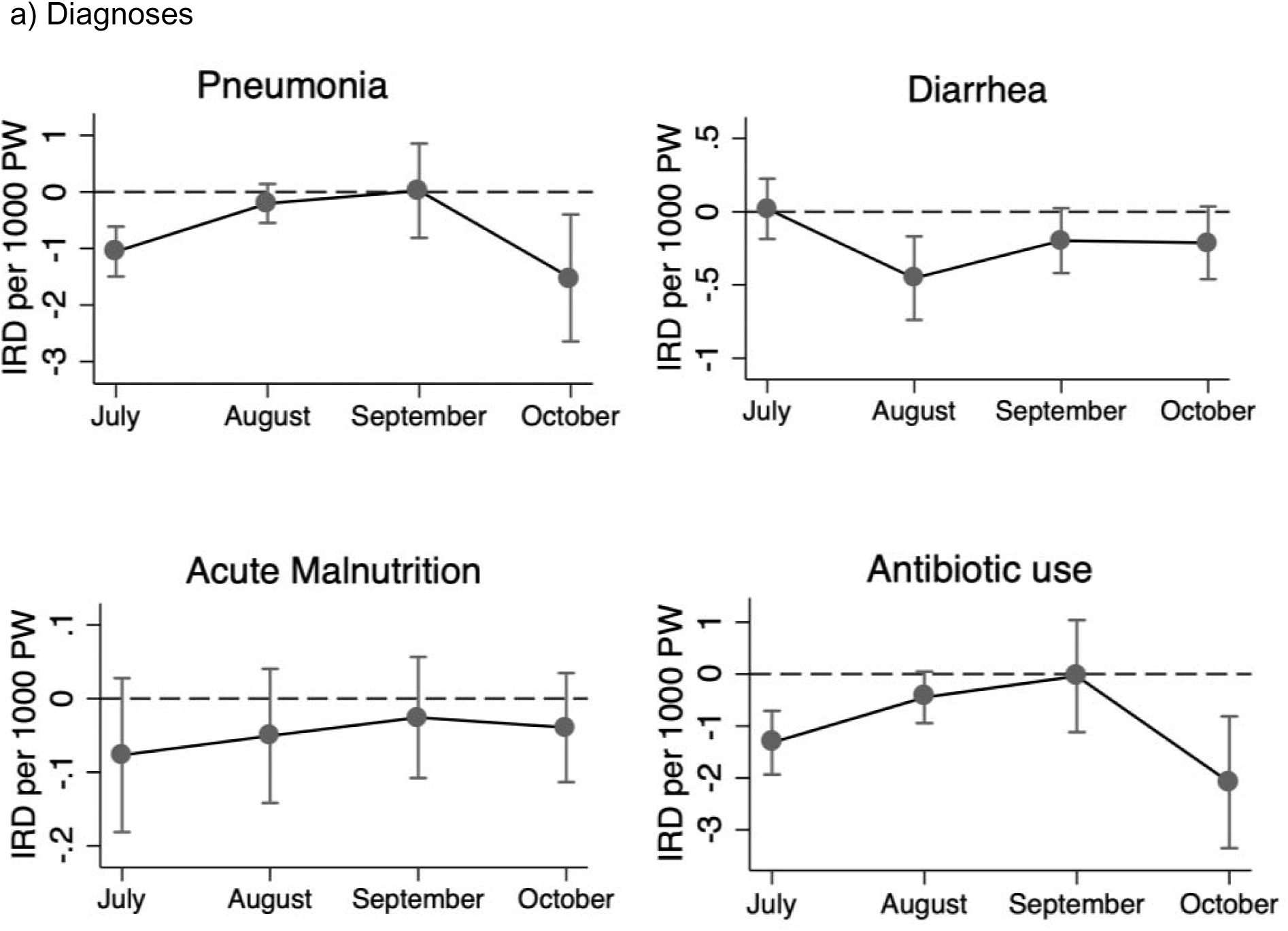
– Difference in Incidence Rates of a) Diagnoses and b) Treatment During SMC Administration vs. Pos-Administration Weeks, by Month of Administration. Note: P-values from the global test of interaction of SMC and month of administration were 0.000, 0.047, and 0.654 for pneumonia, diarrhea, and acute malnutrition, and 0.002 for antibiotic prescription rates

For the control outcomes, the reduction in rates following SMC was also highest in October for both malaria incidence and antimalarial prescription rates, while no significant variation was observed for injury (Supplementary Figure 2).

Although the cycles with the highest reduction varied slightly by outcome, the last round (October) was the period during which the reductions following SMC were seen for most outcomes.

In the sensitivity analyses examining changes in diagnoses and prescription rates in each of the three weeks following administration, rates of diarrhea, acute malnutrition, and antibiotic use declined most in the second week post-administration, while pneumonia rates declined most in the third week post-administration (Supplementary table 2). Rates of the control outcomes, malaria and antimalarial prescription rates also declined most in the second week post-administration.

## Discussion

We evaluated the short-term changes in non-malarial outcomes following SMC administration under routine program conditions in Burkina Faso. We found modest declines in rates of pneumonia, diarrhea and acute malnutrition in the post-SMC period. Additionally, there was a small reduction in antibiotic prescription rates. The change in rates of pneumonia, diarrhea, acute malnutrition and antibiotic prescription following SMC varied by the month of administration, with its overall benefits appearing more pronounced in October (after the final round of administration).

The modest reductions in non-malarial outcomes following SMC may be partially explained by the antimicrobial properties of sulfadoxine-pyrimethamine (SP). In a previous study, SP had a clinically significant effect on concurrent infectious diseases in pregnant women and has demonstrated antibacterial effects in vitro, particularly against pathogens such as *Staphylococcus aureus* and *Streptococcus pneumoniae*.^13^ This suggest that sulfadoxine-pyrimethamine may have an effect against some bacterial pathogens, potentially contributing directly to reductions in pneumonia and diarrhea.

SMC may also indirectly lower susceptibility to other infections by reducing malaria burden. Malaria control strategies may contribute to a reduction in other infections by reducing malaria-related immunosuppression.^33^ Malaria can weaken the immune system because repeated or long-term infections can lead to a dysregulated immune response, making the body less effective at fighting off new or other types of infections.^33^ Previous evidence shows that children with recent or acute malaria are at an increased risk of bacterial infections, with those experiencing both malaria and invasive bacterial infections facing a substantially higher risk of mortality.^34^

Malaria can also exacerbate malnutrition, which in turn increases risk of infections, illness and mortality in children.^35^ A previous study showed that children who received chemoprophylaxis with chloroquine had fewer episodes of severe malnutrition compared to the control group.^36^ Our findings also show modest declines in acute malnutrition following SMC. Therefore, the reduction in malaria burden following SMC could lead to improvements in immunocompetence, malnutrition and overall health, further reducing the risk of other infections or concurrent illnesses.

These indirect health benefits may contribute to reductions in mortality associated with SMC administration. Previous studies have found that SMC reduces all-cause hospitalization and death,^10,11^ although it is not clear whether its effect on all-cause mortality is due primarily to its impact on severe malaria or if it also reduces mortality from other infections. Our findings suggest that SMC’s impact on non-malaria infections may contribute to its previously demonstrated effect on all-cause mortality,^11^ as rates of the most common non-malaria illnesses appear to lower following SMC administration. Future studies should further investigate its role in reducing mortality from non-malarial causes to better understand its broader effects on all-cause mortality in children and its potential integration with other health interventions.

The reductions in antibiotic use following SMC administration may be due to a decrease in febrile illnesses related to malaria, as well as a reduction in non-malarial infections. A previous study assessing indication for antibiotic prescriptions in health facilities in rural Burkina Faso found that 5% of all antibiotic prescriptions were for malaria diagnoses.^23^ Overall, 7.3% of malaria diagnoses received an antibiotic prescription while 97% of pneumonia diagnoses and 92% of dysentery diagnoses resulted in an antibiotic prescription.^23^ While broad-spectrum antibiotic treatment is recommended for children with severe malaria,^37^ some of the antibiotic prescriptions for malaria may be unnecessary or the result of misdiagnosis, as pneumonia and malaria can present with similar clinical signs, including fever.^23,38^

The lower rate of antibiotic prescriptions following SMC may therefore reflect a reduced need for antibiotics, indicating a decrease in the overall burden of infections. This, in turn, could help lower pressure on healthcare systems, particularly in facilities serving multiple communities. A study on the healthcare burden in Burkina Faso showed that infectious illnesses not only strain healthcare systems but also significantly impact households, forcing families to deplete savings, sell livestock or reduce consumption to afford treatment.^39^ Therefore, reducing infections and improving overall health could potentially help ease the burden on health systems and lower treatment costs for families.

Additionally, the reduction in antibiotic prescriptions following SMC could have implications for antibiotic resistance. Estimates of the global burden of bacterial antimicrobial resistance (AMR) indicate that AMR is a major issue in sub-Saharan Africa.^40,41^ A study examining antibiotic prescription rates following universal distribution of long-lasting insecticidal nets (LLIN) and indoor residual spraying of insecticide (IRS) also observed a reduction in antibiotic use and highlighted the potential for effective malaria interventions to help mitigate antibiotic resistance resulting from routine use of antibiotics.^17^ Therefore, there is potential for SMC administration to help reduce subsequent routine antibiotic use at the population level.

The relatively higher benefits of SMC observed in October for pneumonia and antibiotic prescriptions, may be partly due to the seasonality of infections. Rates for these outcomes were higher during the last round of SMC compared to prior rounds (Figure 2). Consequently, the observed benefit of SMC in October may appear more pronounced because the baseline (pre-administration) rates were higher, leading to a larger reduction in infection burden and subsequent treatment prescriptions. A previous study in Togo also found that the greatest reduction in malaria compared to the first round was during the peak periods, in the last round of SMC in October.^18^ Similarly, diarrhea rates particularly in 2020, were slightly higher around the second round of SMC, during which the largest reduction in diarrhea rates following SMC was observed. Although antibiotic prescription rates were slightly higher, the trends mirrored those of pneumonia diagnoses (Figure 2), as the majority of pediatric antibiotic prescriptions in this area of Burkina Faso are related to pneumonia.^23^

This study has some limitations. First, although we used negative and positive control outcomes, we cannot fully rule out unmeasured or residual confounding. SMC campaigns which include community sensitization^42^ may temporarily strengthen health-seeking behaviors and increase adherence to preventive practices,^43^ such as increased bednet use and improved sanitation and hygiene, which may affect the risk of other infections. Furthermore, these indirect benefits of SMC were assessed in a setting where mass distribution of azithromycin was provided to some communities as part of a cluster-randomized trial. Additionally, the years included in the analyses (2020 and 2021) were during the COVID-19 pandemic, which may have affected clinic visits or regular patterns of care-seeking.

Therefore, these findings may have limited generalizability to other periods or areas. Additionally, it is not clear whether non-malarial outcomes, such as pneumonia, diarrhea, and acute malnutrition, occurred independently or if some were concurrent with malaria. This distinction may influence how SMC affects the rates of these other infections in the weeks following administration. Future studies should evaluate this aspect to better understand the full impact of SMC on co-infections and overall health outcomes.

Second, potential misclassification in diagnosing cases could affect our weekly count estimates. Additionally, surveillance data may be subject to underreporting or inconsistencies in data collection. Despite these limitations, it offers large-scale, real-time insights that are helpful for monitoring disease trends in populations and evaluating the need for and impact of interventions.^44^ Lastly, the variability in indirect health benefits over the months may partly reflect chance findings, as only two years of data were available for analysis. Any differences observed could also be mixed with seasonal changes in these outcomes that could mask effects during earlier administrations or be partially due to the waning of the seasonal epidemic of infections (e.g., respiratory pathogens) that could appear as enhanced benefit during particular cycles. Therefore, these results should be interpreted cautiously and confirmed by a study with a longer time frame with multiple years of data, which could provide a more accurate assessment of differences in effects across months and cycles.

## Conclusion

We found modest reductions in non-malarial infectious outcomes following SMC administration, with reductions more pronounced in the last month of administration. Our findings suggest that SMC may have additional benefits beyond malaria prevention, particularly in contributing to reductions in common infectious diseases among children and routine antibiotic use. These findings indicate that SMC could have a broader utility in improving health and potentially lowering pressure on healthcare systems although further investigation is needed to fully assess its broader effects on health outcomes.

## Declarations

### Funding

The CHAT trial through which the clinic surveillance data was collected was supported by the Gates Foundation (grant number OPP1187628). The conclusions and opinions expressed in this work are those of the authors alone and should not be attributed to the Foundation. Under the grant conditions of the Foundation, a Creative Commons Attribution 4.0 License has already been assigned to the Author Accepted Manuscript version that may arise from this submission. Please note that works submitted as preprints have not undergone a peer review process.

Research reported in this manuscript was also supported by the National Institutes of Health Eunice Kennedy Shriver National Institute of Child Health & Human Development (NIH/NICHD) F31 Award (1F31HD114434-01A1: E.A.G.).

## Conflicts of Interest

None declared

## Ethics approval and consent to participate

The Randomized controlled trial from which the data was obtained was reviewed and approved by the Institutional Review Boards at the University of California, San Francisco and the Comité National d’Ethique pour la Recherche (National Ethics Committee of Burkina Faso) in Ouagadougou, Burkina Faso. Written informed consent was obtained from the caregiver of each participant.

## Data Availability

The datasets analyzed during the current study are not yet publicly available but will be made accessible through the Open Science Framework (OSF) repository upon finalization. The data will be shared in accordance with relevant open data sharing guidelines. A link to the repository will be provided once the data is available.

## Consent for publication

Not Applicable

## Author Contributions

Elisabeth A. Gebreegziabher: Conceptualization, Methodology, Software, Formal analysis, Investigation, Visualization, Writing – original draft, Writing – review & editing.

Mamadou Ouattara, Mamadou Bountogo, Boubacar Coulibaly, Thierry Ouedraogo, Elodie Lebas: Project administration, Supervision, Resources, Writing – review & editing.

Valentin Boudo, Huiyu Hu: Data curation, Writing – review & editing.

David V. Glidden, Benjamin F. Arnold: Conceptualization, Statistical analysis, Writing – review & editing.

Kieran S. O’Brien: Writing – review & editing.

Michelle S. Hsiang: Conceptualization, Methodology, Writing – review & editing.

Ali Sié, Thomas M. Lietman, Catherine E. Oldenburg: Conceptualization, Funding acquisition, Project administration, Supervision, Writing – review & editing.

## Author contact information

Elisabeth Gebreegziabher, Mamadou Ouattara, Mamadou Bountogo, Boubacar Coulibaly, Valentin Boudo, Thierry Ouedraogo, Elodie Lebas, Huiyu Hu, Kieran S. O’Brien, Michelle S. Hsiang, David V. Glidden, Benjamin F. Arnold, Thomas M. Lietman, Ali Sié, Catherine E. Oldenburg

## Tables and Figures

**Supplementary Table 1.** – Changes in Diagnoses and Treatment rates following SMC by Month of SMC Administration and Overall (IRR, 95% CI)

| Outcome | Overall | July | August | September | October | P value for global test of interaction |
| --- | --- | --- | --- | --- | --- | --- |
| Pneumonia | 0.86 (0.79 to 0.93) | 0.66 (0.57 to 0.74) | 0.92 (0.80 to 1.05) | 1.0 (0.83 to 1.18) | 0.81 (0.69 to 0.92) | 0.000 |
| Diarrhea | 0.83 (0.74 to 0.93) | 1.02 (0.82 to 1.22) | 0.74 (0.61 to 0.88) | 0.85 (0.70 to 1.0) | 0.82 (0.67 to 0.97) | 0.047 |
| Acute malnutrition | 0.71 (0.51 to 0.97) | 0.49 (0.03 to 0.94) | 0.69 (0.23 to 1.14) | 0.81 (0.29 to 1.34) | 0.80 (0.47 to 1.13) | 0.654 |
| Malaria<br>(positive control) | 0.62 (0.56 to 0.69) | 0.90 (0.73 to 1.07) | 0.59 (0.52 to 0.67) | 0.54 (0.47 to 0.61) | 0.67 (0.58 to 0.75) | 0.000 |
| Injury<br>(negative control) | 0.89 (0.67 to 1.19) | 0.92 (0.46 to 1.37) | 1.03 (0.57 to 1.48) | 0.80 (0.44 to 1.16) | 0.82 (0.43 to 1.20) | 0.861 |
| Antibiotic prescription | 0.88 (0.83 to 0.94) | 0.78 (0.70 to 0.86) | 0.92 (0.84 to 1.0) | 1.0 (0.86 to 1.13) | 0.81 (0.72 to 0.91) | 0.002 |
| Antimalarial<br>prescription<br>(positive control) | 0.63 (0.56 to 0.69) | 0.90 (0.72 to 1.07) | 0.60 (0.53 to 0.68) | 0.54 (0.47 to 0.61) | 0.67 (0.58 to 0.76) | 0.000 |

**Supplementary Table 2.** – Incidence Rates, Rate Ratios, and Rate Differences of Outcomes During SMC Administration Weeks and the First, Second-, and Third-Weeks Post-Administration.

| **Weeks post SMC administration** | **Incidence rate per 1,000 person-week (95%CI)** | **Incidence rate ratio (95%CI)** | **Incidence Rate Difference per 1,000 person-week (95%CI)** |
| --- | --- | --- | --- |
| Malaria (positive control) |  |  |  |
| Admin week | 9.7 (8 to 11.4) | Ref | Ref |
| 1 | 6 (4.9 to 7.1) | 0.61 (0.55 to 0.68) | -3.8 (-4.8 to -2.8) |
| 2 | 5.8 (4.9 to 6.8) | 0.6 (0.53 to 0.68) | -3.9 (-5.1 to -2.7) |
| 3 | 6.4 (5.4 to 7.3) | 0.65 (0.59 to 0.73) | -3.4 (-4.5 to -2.3) |
| Injury (negative control) |  |  |  |
| Admin week | 0.17 (0.12 to 0.23) | Ref | Ref |
| 1 | 0.14 (0.11 to 0.17) | 0.81 (0.58 to 1.13) | -0.03 (-0.09 to 0.02) |
| 2 | 0.17 (0.13 to 0.21) | 0.98 (0.73 to 1.31) | 0.0 (-0.06 to 0.05) |
| 3 | 0.15 (0.11 to 0.2) | 0.89 (0.63 to 1.26) | -0.02 (-0.08 to 0.04) |
| Pneumonia |  |  |  |
| Admin week | 4.7 (3.8 to 5.7) | Ref | Ref |
| 1 | 4.3 (3.5 to 5) | 0.9 (0.83 to 0.97) | -0.5 (-0.9 to -0.1) |
| 2 | 4.1 (3.2 to 4.9) | 0.86 (0.78 to 0.95) | -0.7 (-1.1 to -0.2) |
| 3 | 3.9 (3.1 to 4.6) | 0.82 (0.74 to 0.90) | -0.9 (-1.3 to -0.4) |
| Diarrhea |  |  |  |
| Admin week | 1.4 (1 to 1.7) | Ref | Ref |
| 1 | 1.2 (0.9 to 1.5) | 0.87 (0.77 to 0.98) | -0.2 (-0.3 to 0) |
| 2 | 1 (0.8 to 1.3) | 0.77 (0.66 to 0.90) | -0.3 (-0.5 to -0.1) |
| 3 | 1.2 (0.9 to 1.5) | 0.86 (0.76 to 0.97) | -0.2 (-0.3 to 0) |
| Acute Malnutrition |  |  |  |
| Admin week | 0.17 (0.09 to 0.24) | Ref | Ref |
| 1 | 0.12 (0.05 to 0.2) | 0.74 (0.49 to 1.11) | -0.04 (-0.1 to 0.01) |
| 2 | 0.1 (0.04 to 0.17) | 0.63 (0.42 to 0.94) | -0.06 (-0.11 to -0.01) |
| 3 | 0.13 (0.06 to 0.19) | 0.76 (0.54 to 1.06) | -0.04 (-0.09 to 0.01) |
| Antimalarial prescription (positive control) |  |  |  |
| Admin week | 9.7 (7.9 to 11.4) | Ref | Ref |
| 1 | 6 (4.8 to 7.1) | 0.62 (0.56 to 0.69) | -3.7 (-4.7 to -2.7) |
| 2 | 5.8 (4.8 to 6.8) | 0.60 (0.53 to 0.68) | -3.9 (-5.1 to -2.7) |
| 3 | 6.4 (5.4 to 7.4) | 0.66 (0.59 to 0.74) | -3.3 (-4.4 to -2.2) |
| Antibiotic prescription |  |  |  |
| Admin week | 7.9 (6.6 to 9.2) | Ref | Ref |
| 1 | 7.1 (5.8 to 8.3) | 0.90 (0.84 to 0.96) | -0.82 (-1.33 to -0.31) |
| 2 | 6.9 (5.7 to 8) | 0.87 (0.81 to 0.93) | -1.02 (-1.57 to -0.46) |
| 3 | 6.9 (5.7 to 8.1) | 0.87 (0.8 to 0.95) | -0.99 (-1.61 to -0.36) |

**Supplementary Figure 1.**
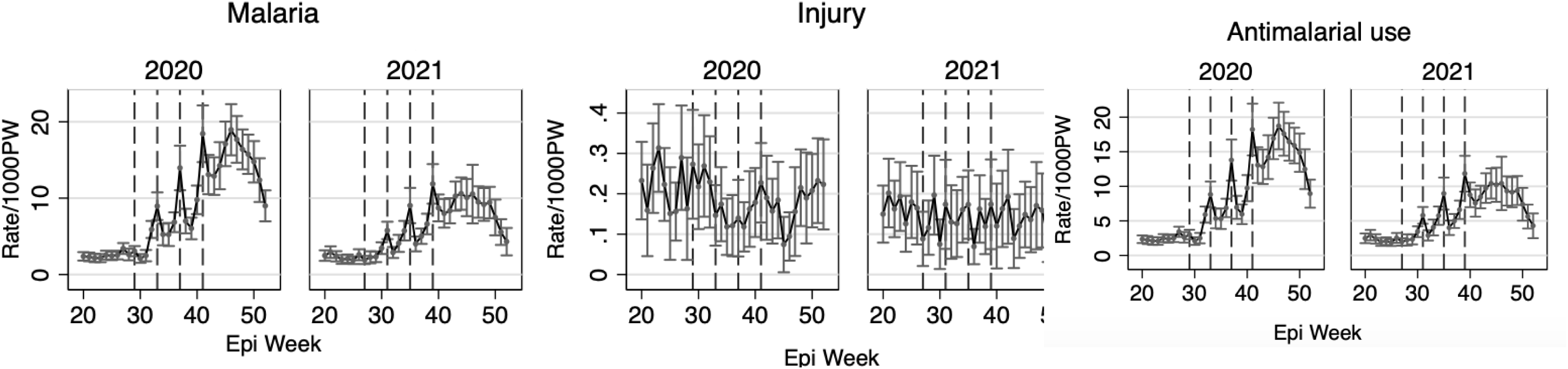
– Diagnoses Rates and Treatment Prescription Rates for Control Outcomes in 2020 and 2021 in the Context of SMC Administration.

**Supplementary Figure 2.**
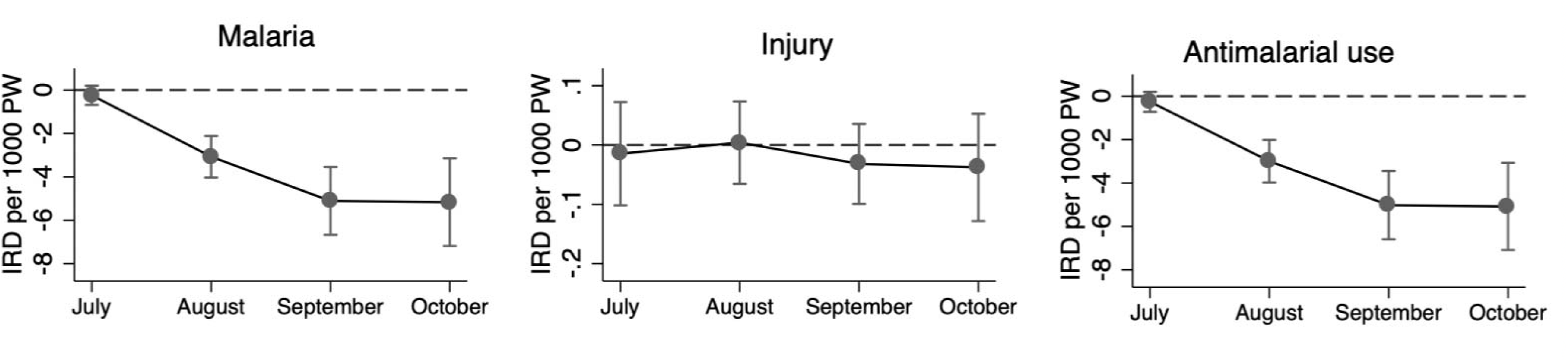
– Difference in Incidence Rates of Diagnoses and Treatment During SMC Administration vs. Post-Administration Weeks, by Month of Administration for Control Outcomes. Note: P-values from the global test of interaction between SMC and month of administration were 0.000 and 0.861 for malaria and injury, respectively, and 0.001 for antimalarial prescription rates.

## Legends

### Tables

**Table 1**-Diagnoses and treatment rates in CSPS/Health Posts in Nouna, Burkina Faso included in the Analysis (n = 51), 2020 and 2021

**Table 2**-Incidence rate, rate ratio and rate difference of outcomes on SMC administration weeks and the three weeks post administration

### Figures

**Figure 1**-Conceptual Framework for SMC, Non-Malarial Outcomes, and Antibiotic Prescription Rates

**Figure 2**-Diagnoses Rates and Treatment Prescription Rates in 2020 and 2021 in the Context of SMC Administration.

**Figure 3**-Difference in Incidence Rates of a) Diagnoses and b) Treatment During SMC Administration vs. Post-Administration Weeks, by Month of Administration

### Supplementary material

**Supplementary Table 1**-Changes in Diagnoses and Treatment rates following SMC by Month of SMC Administration and Overall (IRR, 95% CI)

**Supplementary Table 2**-Incidence Rates, Rate Ratios, and Rate Differences of Outcomes During SMC Administration Weeks and the First, Second-, and Third-Weeks Post-Administration

**Supplementary Figure 1**-Diagnoses Rates and Treatment Prescription Rates for Control Outcomes in 2020 and 2021 in the Context of SMC Administration

**Supplementary Figure 2**-Difference in Incidence Rates of Diagnoses and Treatments During SMC Administration vs. Post-Administration Weeks, by Month of Administration for Control Outcomes

## Notes

### Competing Interest Statement

The authors have declared no competing interest.

### Author Declarations

The study was reviewed and approved by the Institutional Review Boards at the University of California, San Francisco and the Comite National d Ethique pour la Recherche (National Ethics Committee of Burkina Faso) in Ouagadougou, Burkina Faso.

## References

1. Walker CLF, Rudan I, Liu L, et al. Global burden of childhood pneumonia and diarrhoea. Lancet. Apr 20 2013;381(9875):1405–1416. doi:10.1016/s0140-6736(13)60222-6

2. Ndiaye M, Sylla K, Sow D, et al. Potential Impact of Seasonal Malaria Chemoprevention on the Acquisition of Antibodies Against Glutamate-Rich Protein and Apical Membrane Antigen 1 in Children Living in Southern Senegal. Am J Trop Med Hyg. Oct 2015;93(4):798–800. doi:10.4269/ajtmh.14-0808

3. World Health Organization. Malaria Prevention Works, Let’s Close the Gap. Geneva, Switzerland: World Health Organization; 2017: 127–129 Retrieved February 27, 2023

4. Seasonal malaria chemoprevention with sulfadoxine–pyrimethamine plus amodiaquine in children: a field guide. Global Malaria Programme; World Health Organization; Second edition; ISBN: 978-92-4-007369-2. May 2023;

5. Alliance S. 53 million children living in 18 countries covered with Seasonal Malaria Chemoprevention in 2023. Retrieved March 4, 2025 April 5, 2024;

6. WHO guidelines for malaria. Geneva: World Health Organization; 2023 (WHO/UCN/GMP/ 202301 Rev1) License: CC BY-NC-SA 30 IGO). 16 October 2023;

7. Dicko A, Diallo AI, Tembine I, et al. Intermittent preventive treatment of malaria provides substantial protection against malaria in children already protected by an insecticide-treated bednet in Mali: a randomised, double-blind, placebo-controlled trial. PLoS Med. Feb 1 2011;8(2):e1000407. doi:10.1371/journal.pmed.1000407

8. Khan J, Suau Sans M, Okot F, et al. A quasi-experimental study to estimate effectiveness of seasonal malaria chemoprevention in Aweil South County in Northern Bahr El Ghazal, South Sudan. Malaria Journal. 2024/01/24 2024;23(1):33. doi:10.1186/s12936-024-04853-x

9. Partnership A-S. Effectiveness of seasonal malaria chemoprevention at scale in west and central Africa: an observational study. Lancet. Dec 5 2020;396(10265):1829–1840. doi:10.1016/s0140-6736(20)32227-3

10. Wilson AL. A systematic review and meta-analysis of the efficacy and safety of intermittent preventive treatment of malaria in children (IPTc). PLoS One. Feb 14 2011;6(2):e16976. doi:10.1371/journal.pone.0016976

11. Issiaka D, Barry A, Traore T, et al. Impact of seasonal malaria chemoprevention on hospital admissions and mortality in children under 5 years of age in Ouelessebougou, Mali. Malar J. Mar 3 2020;19(1):103. doi:10.1186/s12936-020-03175-y

12. Chandramohan D, Dicko A, Zongo I, et al. Effect of Adding Azithromycin to Seasonal Malaria Chemoprevention. N Engl J Med. Jun 6 2019;380(23):2197–2206. doi:10.1056/NEJMoa1811400

13. Capan M, Mombo-Ngoma G, Makristathis A, Ramharter M. Anti-bacterial activity of intermittent preventive treatment of malaria in pregnancy: comparative in vitro study of sulphadoxine-pyrimethamine, mefloquine, and azithromycin. Malar J. Oct 29 2010;9:303. doi:10.1186/1475-2875-9-303

14. Faure E. Malarial pathocoenosis: beneficial and deleterious interactions between malaria and other human diseases. Front Physiol. 2014;5:441. doi:10.3389/fphys.2014.00441

15. Smithson P, Florey L, Salgado SR, et al. Impact of Malaria Control on Mortality and Anemia among Tanzanian Children Less than Five Years of Age, 1999-2010. PLoS One. 2015;10(11):e0141112. doi:10.1371/journal.pone.0141112

16. Kang H, Kreuels B, Adjei O, Krumkamp R, May J, Small DS. The causal effect of malaria on stunting: a Mendelian randomization and matching approach. Int J Epidemiol. Oct 2013;42(5):1390–8. doi:10.1093/ije/dyt116

17. Krezanoski PJ, Roh ME, Rek J, et al. Marked reduction in antibiotic usage following intensive malaria control in a cohort of Ugandan children. BMC Med. Nov 30 2021;19(1):294. doi:10.1186/s12916-021-02167-2

18. Bakai TA, Thomas A, Iwaz J, et al. Effectiveness of seasonal malaria chemoprevention in three regions of Togo: a population-based longitudinal study from 2013 to 2020. Malaria Journal. 2022/12/31 2022;21(1):400. doi:10.1186/s12936-022-04434-w

19. Sié A, Louis VR, Gbangou A, et al. The Health and Demographic Surveillance System (HDSS) in Nouna, Burkina Faso, 1993-2007. Glob Health Action. Sep 14 2010;3 doi:10.3402/gha.v3i0.5284

20. Sié A, Ouattara M, Bountogo M, et al. A double-masked placebo-controlled trial of azithromycin to prevent child mortality in Burkina Faso, West Africa: Community Health with Azithromycin Trial (CHAT) study protocol. Trials. Dec 4 2019;20(1):675. doi:10.1186/s13063-019-3855-9

21. Oldenburg CE, Sié A, Ouattara M, et al. Distance to primary care facilities and healthcare utilization for preschool children in rural northwestern Burkina Faso: results from a surveillance cohort. BMC Health Serv Res. Mar 9 2021;21(1):212. doi:10.1186/s12913-021-06226-5

22. Oldenburg CE, Ouattara M, Bountogo M, et al. Mass Azithromycin Distribution to Prevent Child Mortality in Burkina Faso: The CHAT Randomized Clinical Trial. JAMA. 2024;331(6):482–490. doi:10.1001/jama.2023.27393

23. Sié A, Ouattara M, Bountogo M, et al. Indication for Antibiotic Prescription Among Children Attending Primary Healthcare Services in Rural Burkina Faso. Clinical Infectious Diseases. 2021;73(7):1288–1291. doi:10.1093/cid/ciab471

24. Arias JR. EPI Week Calendars 2008-2024. Central MassMosquito Control Project.

25. Harris AD, McGregor JC, Perencevich EN, et al. The use and interpretation of quasi-experimental studies in medical informatics. J Am Med Inform Assoc. Jan-Feb 2006;13(1):16–23. doi:10.1197/jamia.M1749

26. Cairns ME, Sagara I, Zongo I, et al. Evaluation of seasonal malaria chemoprevention in two areas of intense seasonal malaria transmission: Secondary analysis of a household-randomised, placebo-controlled trial in Houndé District, Burkina Faso and Bougouni District, Mali. PLoS Med. Aug 2020;17(8):e1003214. doi:10.1371/journal.pmed.1003214

27. Gebreegziabher EO, Mamadou; Bountogo, Mamadou et al. Trends in Uncomplicated and Severe Malaria following Seasonal Malaria Chemoprevention Administration in Nouna, Burkina Faso. PREPRINT available at Research Square. July 2024;

28. Lopez Bernal J, Cummins S, Gasparrini A. The use of controls in interrupted time series studies of public health interventions. International Journal of Epidemiology. 2018;47(6):2082–2093. doi:10.1093/ije/dyy135

29. Arnold BF, Ercumen A. Negative Control Outcomes: A Tool to Detect Bias in Randomized Trials. Jama. Dec 27 2016;316(24):2597–2598. doi:10.1001/jama.2016.17700

30. Lipsitch M, Tchetgen Tchetgen E, Cohen T. Negative controls: a tool for detecting confounding and bias in observational studies. Epidemiology. May 2010;21(3):383–8. doi:10.1097/EDE.0b013e3181d61eeb

31. Clarke D, Romano JP, Wolf M. The Romano–Wolf multiple-hypothesis correction in Stata. The Stata Journal. 2020/12/01 2020;20(4):812–843. doi:10.1177/1536867X20976314

32. Cairns M, Carneiro I, Milligan P, et al. Duration of protection against malaria and anaemia provided by intermittent preventive treatment in infants in Navrongo, Ghana. PLoS One. May 21 2008;3(5):e2227. doi:10.1371/journal.pone.0002227

33. Calle CL, Mordmüller B, Singh A. Immunosuppression in Malaria: Do Plasmodium falciparum Parasites Hijack the Host? Pathogens. Oct 3 2021;10(10)doi:10.3390/pathogens10101277

34. Church J, Maitland K. Invasive bacterial co-infection in African children with Plasmodium falciparum malaria: a systematic review. BMC Med. Feb 19 2014;12:31. doi:10.1186/1741-7015-12-31

35. Child Malnutrition. Unicef datauniceforg/topic/nutrition/malnutrition/May 2023

36. Bradley-Moore AM, Greenwood BM, Bradley AK, Kirkwood BR, Gilles HM. Malaria chemoprophylaxis with chloroquine in young Nigerian children. Annals of Tropical Medicine & Parasitology. 1985/01/01 1985;79(6):575–584. doi:10.1080/00034983.1985.11811964

37. World Health O. Management of severe malaria: a practical handbook. 3rd ed. World Health Organization; 2012.

38. Kaboré B, Post A, Lompo P, et al. Aetiology of acute febrile illness in children in a high malaria transmission area in West Africa. Clinical Microbiology and Infection. 2021/04/01/ 2021;27(4):590–596. doi:doi: 10.1016/j.cmi.2020.05.029

39. Yaya Bocoum F, Grimm M, Hartwig R. The health care burden in rural Burkina Faso: Consequences and implications for insurance design. SSM Popul Health. Dec 2018;6:309–316. doi:10.1016/j.ssmph.2018.10.012

40. Laxminarayan R. The overlooked pandemic of antimicrobial resistance. The Lancet. 2022;399(10325):606–607. doi:10.1016/S0140-6736(22)00087-3

41. Murray CJL, Ikuta KS, Sharara F, et al. Global burden of bacterial antimicrobial resistance in 2019: a systematic analysis. The Lancet. 2022;399(10325):629–655. doi:10.1016/S0140-6736(21)02724-0

42. Preventing Childhood Malaria through Seasonal Malaria Chemoprevention (SMC): Three Big Lessons. PMI Impact Malaria USAID; 2019 Retrieved January 23, 2025 blog. 2019.

43. Onyinyechi OM, Mohd Nazan AIN, Ismail S. Effectiveness of health education interventions to improve malaria knowledge and insecticide-treated nets usage among populations of sub-Saharan Africa: systematic review and meta-analysis. Front Public Health. 2023;11:1217052. doi:10.3389/fpubh.2023.1217052

44. Nsubuga P WM, Thacker SB, et al. Public Health Surveillance: A Tool for Targeting and Monitoring Interventions. Disease Control Priorities in Developing Countries 2nd edition Washington (DC): The International Bank for Reconstruction and Development / The World Bank; 2006 Chapter 53 Co-published by Oxford University Press, New York.

